# Flexible Bayesian estimation of incubation times

**DOI:** 10.1101/2023.08.07.23293752

**Authors:** Oswaldo Gressani, Andrea Torneri, Niel Hens, Christel Faes

**Affiliations:** Interuniversity Institute for Biostatistics and statistical Bioinformatics (I-BioStat), Data Science Institute, Hasselt University, Hasselt, Belgium; Centre for Health Economics Research and Modelling Infectious Diseases, Vaxinfectio, University of Antwerp, Antwerp, Belgium

**Keywords:** Incubation period, Laplace approximation, Bayesian P-splines, MCMC

## Abstract

**Motivation:** The incubation period is of paramount importance in infectious disease epidemiology as it informs about the transmission potential of a pathogenic organism and helps to plan public health strategies to keep an epidemic outbreak under control. Estimation of the incubation period distribution from reported exposure times and symptom onset times is challenging as the underlying data is coarse.

**Methodology:** We develop a new Bayesian methodology using Laplacian-P-splines that provides a semi-parametric estimation of the incubation density based on a Langevinized Gibbs sampler. A finite mixture density smoother informs a set of parametric distributions via moment matching and an information criterion arbitrates between competing candidates.

**Results:** Our method has a natural nest within EpiLPS, a tool originally developed to estimate the time-varying reproduction number. Various simulation scenarios accounting for different levels of data coarseness are considered with encouraging results. Applications to real data on COVID-19, MERS-CoV and Mpox reveal results that are in alignment with what has been obtained in recent studies.

**Conclusion:** The proposed flexible approach is an interesting alternative to classic Bayesian parametric methods for estimation of the incubation distribution.

## 1 Introduction

Statistical methods and their underlying algorithmic implementation play an essential role in infectious disease modeling as they permit to bridge the gap between observed data and estimates of key epidemiologic quantities. The incubation period, defined as the time between infection and symptom onset (Lessler et al., 2009) is pivotal in gauging the epidemic potential of an infectious disease. Having information about the incubation period distribution is helpful for planning optimal quarantine periods to taper off the spread of a contagious disease (Qin et al., 2020). Knowledge of incubation times helps in assessing the transmission potential of an infectious disease (Cheng et al., 2021; Basnarkov et al., 2022) as the incubation period can be used to estimate the reproduction number (i.e., the average number of secondary cases generated by an infector in a fully susceptible population). The incubation period is also of direct interest for case definition (Virlogeux et al., 2016) and to measure the effectiveness of contact tracing. Moroever, it contributes in quantifying the size of an epidemic (Backer et al., 2020) and improves the ecological comprehension of adaptation strategies of a parasite (Nishiura, 2007). The centrality of incubation features in epidemic analyses thus calls for solid methodologies that provide accurate and reliable estimates of the incubation distribution to better understand the transmission dynamics of a pathogen and to reach effective interventional public health strategies.

From a statistical point of view, the main obstacle for inferring the distribution of the incubation period lies in the fact that infection times are almost never exactly observed (Chen et al., 2022), while symptom onset times are more easily observed and reported. This incomplete information set-up pushes towards a more challenging inference approach based on coarse data (Reich et al., 2009), where infection times are only known to lie within a finite time interval. In survival analysis, such a data structure is known as interval-censored data and several approaches have been proposed to estimate the survival function and related quantities from coarsened observations. The paper of Peto (1973) is among the first to propose a modeling attempt under an interval-censoring mechanism, where maximum likelihood estimation is carried out through a constrained Newton-Raphson algorithm and applied to a survey on sexual maturity development. A now popular extension considered by Turnbull (1976) consists in using a kind of expectation-maximization algorithm to build a non-parametric estimate of the cumulative distribution function under a more general form of data incompleteness that includes interval-censoring. These two pioneering papers provided a fertile soil for the development of other frequentist approaches (see e.g. Gómez et al., 2004, 2009). Bayesian methods for interval-censored data are more recent as practical implementations had to wait for the arrival of modern machines that facilitated the use of Markov chain Monte Carlo (MCMC) algorithms to extract information from complex posterior distributions. Sinha and Dey (1997) give a comprehensive review of semi-parametric Bayesian methods for survival data characterizd by interval-censoring among others and the work of Calle and Gómez (2001) presents a non-parametric Bayesian estimator of the survival curve using Gibbs sampling under a Dirichlet process prior.

More directly related to infectious disease epidemiology, the work of Reich et al. (2009) proposes frequentist parametric approaches to estimate the incubation period distribution using the accelerated failure time model with applications to influenza A and RSV. Backer et al. (2020) and Miura et al. (2022) use a Bayesian parametric approach to estimate the incubation period of COVID-19 and of Mpox, respectively. Groeneboom (2021) derives a smooth non-parametric estimator of the incubation time distribution by adding a bandwidth parameter that controls the trade-off between noise and bias and Kreiss and Van Keilegom (2022) propose a semi-parametric method to estimate the incubation period based on Laguerre polynomials.

In this paper, we develop a new Bayesian approach to estimate the incubation period distribution articulated around Laplacian-P-splines (Gressani and Lambert, 2018; Gressani et al., 2022a). Our strategy works in two steps. First, we compute a semi-parametric estimate of the incubation density based on a finite mixture density. The component densities are all modeled by means of penalized cubic B-splines (Eilers and Marx, 1996) but with different data representations. In the particular case of a two-component structure considered here, the first component density is approached through a single interval-censored likelihood, while the second density is approached through a midpoint imputation of the data, i.e. the missing infection time is artificially fixed at the midpoint of the observed incubation interval. Markov chain Monte Carlo with a Langevinized Gibbs sampler is used to construct the flexible semi-parametric incubation density estimate, where the analytically derived gradient and Hessian of the conditional posterior of the spline components are used to speed up the sampling process. Second, the semi-parametric density estimate of the incubation period is used to fit popular parametric distributions that are often used to model the distribution of the incubation period through a moment matching approach and the best fitting model among the semi-parametric and fully parametric candidates is selected through the Bayesian information criterion.

The article is organized as follows. Section 2 gives a detailed account of the methodology and presents the Bayesian model to construct the semi-parametric estimate with Laplacian-P-splines. We also show how our estimate based on the imputation approach directly benefits from the negative binomial model of the EpiLPS architecture (Gressani et al., 2022b). The matching moment approach to fit the parametric distributions is also described here along with a formulation of the chosen information criterion used for model comparison. A complete simulation study is presented in Section 3, where we assess how our methodology performs under varying levels of data coarseness, different target incubation distributions and sample size. In Section 4, we apply our method to data on COVID-19, MERS and Mpox and make a comparison with results obtained from recent studies. Finally, Section 5 concludes with a discussion for future research and limitations of our work. A routine reflecting the proposed methodology to estimate the incubation density has been added to the EpiLPS package (Gressani, 2021) and a dedicated repository (https://github.com/oswaldogressani/Incubation) permits to reproduce the results of the manuscript.

## 2 Methods

### 2.1 Coarsely observed data

The observed symptom onset time for individual *i* is denoted by 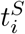 and the (unobserved) infection time is only known to lie within the closed exposure interval 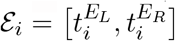, where 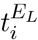 and 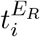 denote the left and right bound, respectively, of the infecting exposure time. Without loss of generality, we work from a continuous time perspective and assume that 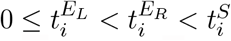 and that symptom onset times are finite. The incubation time is thus at least 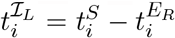 and at most 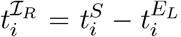, so that the observed data at the resolution of individual *i* is given by the bounds of the incubation period 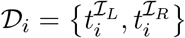 and the information of an observable set of size *n* is thus 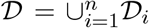. Figure 1 gives a graphical illustration of the relation between exposure times, incubation bounds and the symptom onset time for individual *i*.

**Figure 1.**
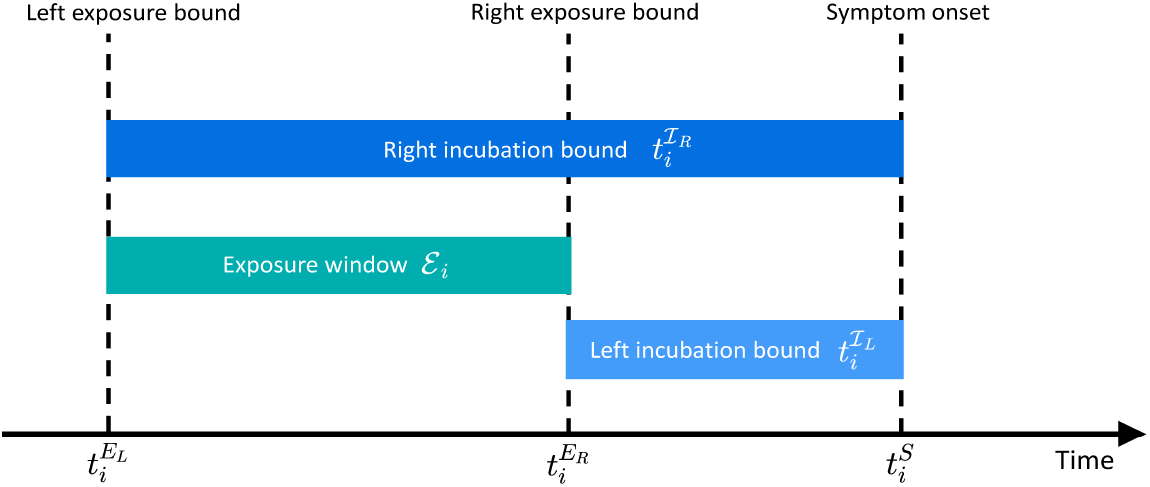
Relation between exposure times, incubation bounds and the symptom onset time.

### 2.2 Semi-parametric model with Bayesian P-splines

Let the incubation time ℐ be a non-negative continuous random variable with probability density function *φ*(*·*), hazard function *h*(*·*) and survival function *S*(*·*). Based on a dataset *𝒟*, we propose to estimate *φ*(*·*) by a two-component mixture density using a semi-parametric (SP) approach based on P-splines. The candidate density estimator at a given time point *t ≥* 0 is denoted by 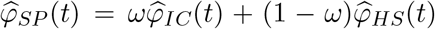, with 0 *≤ ω ≤* 1. The density estimator 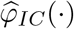 is based on single interval-censored (IC) data as shown in Figure 1, while 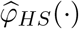 is a density estimator resulting from a histogram smoother assuming a midpoint imputation rule for the infection time in the exposure window *ε*. The next two sections give a detailed outline of the models underlying the latter two component densities.

#### 2.2.1 Flexible density estimation for single interval-censored data

Following Rosenberg (1995), the (log-)hazard of the incubation period is approximated by a linear combination of cubic B-spline basis functions:

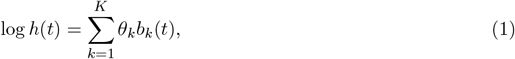

where *b*(*·*) = (*b*_1_(*·*), …, *b*_*K*_(*·*)^⊤^ is a cubic B-spline basis with equidistant knots on the compact time interval *𝒯* =[0, *t*_*u*_] with upper bound *t*_*u*_ and ***θ*** = (*θ*_1_, …, *θ*_*K*_)^⊤^ is the *K*-dimensional latent vector of B-spline amplitudes. While zero is a natural lower bound for the incubation period, there is no natural choice for the upper bound *t*_*u*_. An intuitive candidate would be to fix it at the largest observed right bound of the incubation time, i.e. 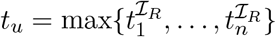, however the latter choice may restrict the B-spline basis to a domain that covers only a small fraction of the domain of the true underlying incubation density *φ*(*·*). As such, we follow Eilers and Marx (2021) and advise to pad *t*_*u*_ to a value that is strictly larger than the largest observed incubation bound. We defer the discussion on the guidelines for a smart padding choice to the real data applications in Section 4. Regarding the number *K* of B-spline basis functions, a default choice in the present setting is *K* = 10, although larger numbers, say *K* = 20 or *K* = 30 may be necessary to capture more flexible patterns (for instance if the underlying incubation density has multiple modes). As noted by Eilers and Marx (2021), there is no fear to choose a “too large” number *K*, as the penalty will act as a counterforce to the induced flexibility.

Using the relation between the survival and hazard functions, we recover:

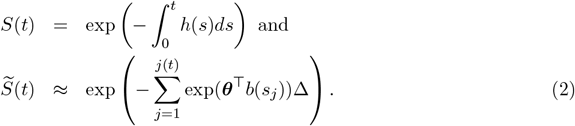

The approximation in (2) is necessary as the integral has no analytic solution. As such, 𝒯 is partitioned into a large number of *J* (e.g. *J* = 300) bins of equal width ∆, where *s*_*j*_ denotes the center of the *j*th bin and *j*(*t*) ∈ {1, …, *J*} is an index function returning the bin number containing *t*. Following Lang and Brezger (2004), a zero-mean Gaussian prior is imposed on the vector of B-spline amplitudes ***θ***|*λ ∼ 𝒩*_dim(***θ***)_ (0, (*λP*)^*−*1^, where *λ >* 0 is the penalty parameter related to the spline model and 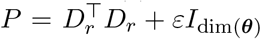 is a square penalty matrix obtained from *− × r*th order difference matrices *D*_*r*_ of dimension (dim(***θ***)− *r*) × dim(***θ***) perturbed by an *ε*-multiple (here *ε* = 10^−6^) to ensure *P* fulfils full rankedness. The Bayesian model is closed by assuming a non-informative Gamma prior on the penalty parameter *λ* ∼ 𝒢(*a*_*λ*_, *b*_*λ*_) with shape *a*_*λ*_ = 10^−4^ and rate *b*_*λ*_ = 10^−4^ (see e.g. Lambert and Eilers, 2005, 2009). The (log-)likelihood of incubation times under single interval-censored data is (Reich et al., 2009):

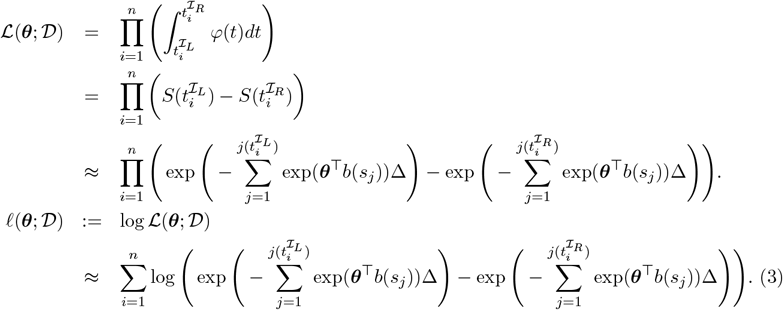

From Bayes’ theorem, one obtains the (log-)conditional posterior density (for a given value of *λ*) of the vector of B-spline coefficients:

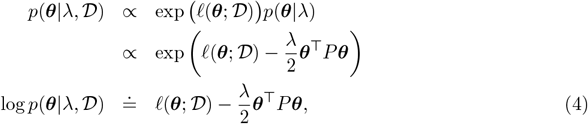

where *∝* and ≐ are symbols used to denote equality up to a multiplicative and additive constant, respectively. The Laplace approximation to the conditional posterior of the B-spline amplitudes is obtained by fitting a (multivariate) Gaussian density around the (unknown) mode ***θ***_*M*_ (*λ*) of *p*(***θ***|*λ, 𝒟*). A Newton-Raphson algorithm involving the gradient and Hessian matrix of log *p*(***θ***|*λ*, 𝒟) is used to approximate the modal value, so that at convergence, one recovers the Laplace approximation 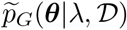 with mean/mode ***θ***^∗^(*λ*) *≈* ***θ***_*M*_ (*λ*) and variance-covariance matrix equal to the inverse of the negative Hessian matrix of log *p*(***θ***❘*λ*, 𝒟) evaluated at ***θ***\*(*λ*) denoted by Σ*(*λ*). To speed-up the mode finding algorithm, we compute the following analytical versions of the gradient and Hessian:

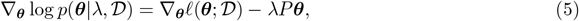

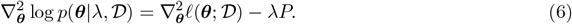

To ease the notation, let us define the following quantities related to the left bound of the incubation period 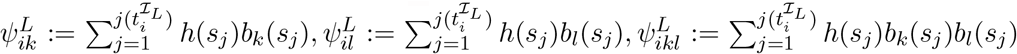 and analogously for the right bound 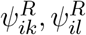 and 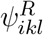. The gradient of the log-likelihood given in (3) is shown to be (see detailed derivations in Appendix S1):

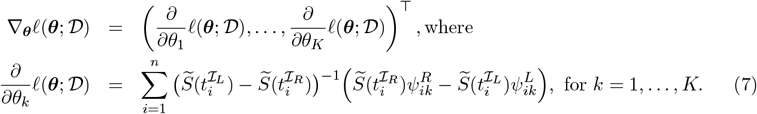

The Hessian matrix of the log-likelihood is (details in Appendix S1):

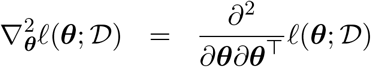, where the entry at the *k*th row and *l*th column is:

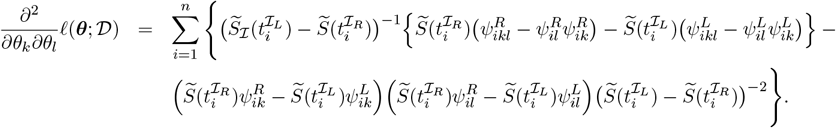

Using the above gradient and Hessian in conjunction with (5) and (6), an iterative algorithm (e.g. Newton-Raphson) can be used to obtain ***θ***^∗^(*λ*) as a proxy for the posterior mode of *p*(***θ***|*λ*, 𝒟). The mode of the Laplace approximation is conditional on the penalty parameter and we therefore need a strategy to calibrate the amount of smoothing. The idea is to use an optimal smoothing approach where the maximum *a posteriori* value of an approximate version of the marginal posterior of *λ* following from Tierney and Kadane (1986) and Rue et al. (2009) is computed. Mathematically, optimal smoothing means 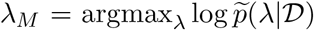, with the following (approximate) posterior distribution for the penalty:

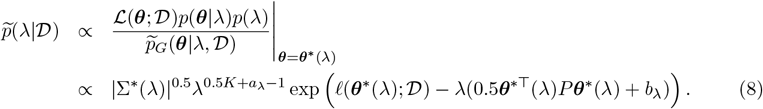

An approximation to *λ*_*M*_ denoted by *λ*^∗^ is found by exploring 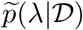 on a linear grid for log_10_(*λ*) and the final resulting Laplace approximation is written by abuse of notation as 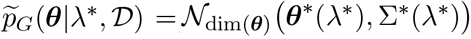.

The mean ***θ***^∗^(*λ*^∗^) and variance-covariance matrix Σ^∗^(*λ*^∗^) are essential quantities to build an efficient MCMC algorithm to sample from the joint posterior of the model parameters *p*(***θ***, *λ*| 𝒟). In fact, we can make use of the Langevinized Gibbs sampler (LGS) developed in Gressani et al. (2022b), where the conditional posterior *p*(***θ***|*λ*, 𝒟) is sampled using a modified Langevin-Hastings algorithm and the conditional posterior of the penalty parameter (*λ*|***θ***, *𝒟*) ∼ 𝒢 (0.5*K*+*a*_*λ*_, 0.5***θ***^⊤^ *P**θ***+*b*_λ_) is sampled in a Gibbs step. The algorithm benefits from adaptive tuning to reach the optimal acceptance rate of 0.57 (Roberts and Rosenthal, 1998). Given {***θ***^(*m*)^ : *m* = 1, …, *M*}, a sample of size *M* of *K*-dimensional vectors ***θ*** obtained frome the LGS algorithm, the point estimate of the *k*th component is taken to be the posterior median of the sample 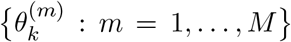 and we denote by 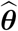 the point estimate of ***θ***. Plugging the latter in the formulas of the hazard in (1) and the survival in (2), we obtain the point estimates 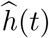 and 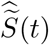 at a given time point *t*. Finally, exploiting the relationship between the density, the hazard and the survival functions, our semi-parametric estimate of the incubation density based on interval-censored data is 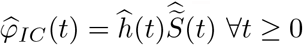.

#### 2.2.2 Flexible density estimation for midpoint imputation

The second component of the mixture density estimator 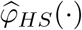 under the semi-parametric approach is obtained through a midpoint imputation technique. Starting from the incubation bounds in *𝒟*, we construct an artificial dataset 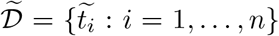, where the infection time of individual *i* is assumed to be located in the middle of the incubation interval, so that the imputed incubation period is:

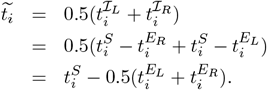

Note that 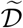 is seen as a random sample from the incubation density *φ*(*·*). From ideas in Eilers and Marx (2010), we construct a histogram on a compact domain 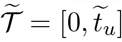 and recommmend to use an upper bound that is at least equal to *t*_*u*_ in Section 2.2.1, i.e. 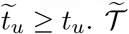 is partitioned in *L* bins with midpoint *x*_*l*_ and width *h* so that the *l*th bin is the half-open interval ℬ_*l*_ = [*x*_*l*_ *− h/*2, *x*_*l*_ + *h/*2) and the last bin is a closed interval. Typically, the histogram smoother is insensitive to the choice of the binwidth (Eilers and Borgdorff, 2007) and it is advocated to use narrow bins (e.g. *h* = 0.05). Another possibility is to use a binwidth *h* determined by a preliminary kernel smoother. The number of imputed incubation periods falling in bin *l* is 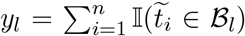, where 𝕝 (*·*) is the indicator function.

The count variable *y*_*l*_ is assumed to follow a negative binomial distribution *y*_*l*_ *∼* NegBin(*μ*_*l*_, *ρ*) with mean *μ*_*l*_ *>* 0 and overdispersion parameter *ρ >* 0. Following the footsteps of Section 2.2.1, we impose a cubic B-spline basis on 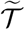 and model the log of the mean counts as 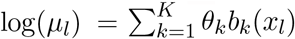. The beauty behind such a formulation is that it allows us to recover exactly the same model as in EpiLPS (Gressani et al., 2022b) to smooth case counts. We thus refer the reader to the latter reference to consult all the equations related to the Laplacian-P-splines approach leading to an estimate of the vector of B-spline coefficients 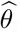. The density estimate resulting from histogram smoothing is then given by: 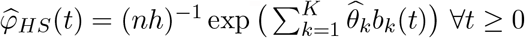 and assuming equal weights *ω* = 0.5, our semi-parametric mixture density estimator for the incubation density *φ*(*t*) at a given time point *t ≥* 0 is 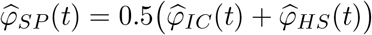.

### 2.3 Parametric fits using moment matching

In some situations it may be advantageous to fit the data by using well-known parametric distributions. Our methodology leaves a door open for this possibility by informing three classic parametric distributions that are usually considered in the estimation of the incubation period, namely the two-parameter lognormal, Gamma and Weibull families. We use *α* and *β* to generically denote the two parameters of the latter families (see Appendix S2 for the detailed parameterization). The moment matching strategy is illustrated in the following pseudo-algorithm:

#### Moment matching algorithm to fit parametric distributions

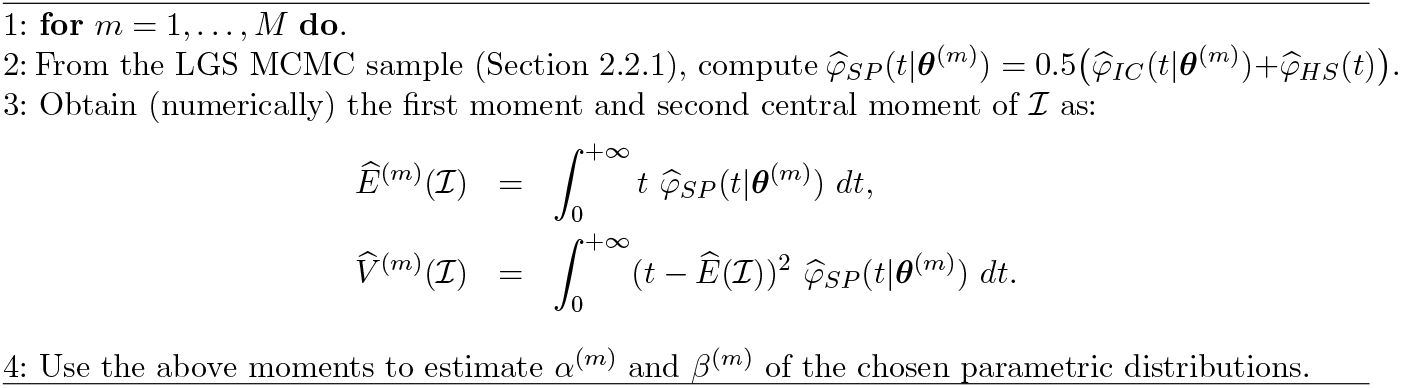

The posterior median of the samples {*α*^(*m*)^ : *m* = 1, …, *M*} and {*β*^(*m*)^ : *m* = 1, …, *M*} denoted by 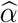 and 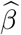, respectively, can be used to construct the lognormal density fit *φ*_*LN*_ (*t*), the Gamma density fit 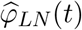 and the Weibull density fit 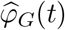. and the Weibull density fit 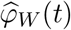 to *φ* (*t*).

To choose between the four candidate density estimates 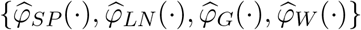, we use the Bayesian information criterion (Schwarz, 1978) computed as 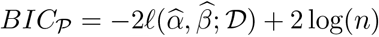 for the parametric fits, i.e. *𝒫* ∈{*LN, G, W*} and 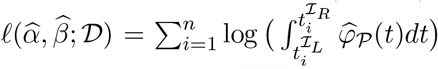 For the semi-parametric fit with P-splines, we use the formula 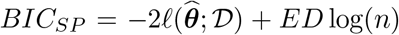, where 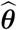 is the estimate of ***θ*** obtained from the LGS algorithm and *ED* is the effective dimension of the model in Section 2.2.1 obtained as follows 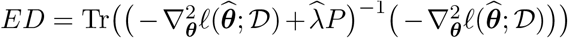, where 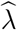 is the median of the MCMC sample for *λ* in the LGS algorithm and Tr(*·*) denotes the trace of a matrix.

## Results

To assess the performance of our methodology, we designed various simulation scenarios with different target incubation densities, data coarseness and sample size. For the incubation density, we use:

- The lognormal density reported in Ferretti et al. (2020) with a mean of 5.5 days and standard deviation of 2.1 days.
- The Weibull density from Backer et al. (2020) with a mean of 6.4 days and standard deviation of 2.3 days.
- An artificial bimodal incubation density constructed as a mixture of two Weibulls with a mean of 7.5 days and standard deviation of 4.6 days.
- A Gamma density from Donnelly et al. (2003) with a mean of 3.8 days and standard deviation of 2.9 days.

We assume two levels of data coarseness with average exposure window *E* equal to one or two days and exposure windows with maximum width of 7 days, reflecting a range that is often observed in practice (see e.g. Yang et al., 2020). For the sample size, we fix *n* = 100 and *n* = 200. From the combination of all these settings, we obtain a total of 4 *×* 2 *×* 2 = 16 scenarios. The features on which we assess the performance are the mean and standard deviation of the incubation period and additional quantiles that are typically of particular interest (e.g. the 5*th* and 95*th* percentiles and the median). We also make a graphical evaluation of the fits by overlaying the density estimates with the target incubation density. Moreover, we are also interested in the performance of the selection process of our methodology, i.e. how many times our modeling approach selects the correct parametric family that corresponds to the incubation distribution used in the data generating mechanism.

For each scenario, we simulate *S* = 1000 datasets and use *K* = 10 B-spline basis functions for all scenarios, except for the bimodal scenario where *K* = 20 to capture the more flexible density pattern. The number of MCMC iterations for the LGS sampler is fixed at *M* = 1000 and the acceptance rate varied in a close neighborhood of the optimal acceptance rate (57%) in all scenarios. Simulations are implemented on an Intel Xeon W-2255 CPU @3.70GHz with 32Go of RAM and it takes approximately one hour for each scenario (and a bit more for the case with *K* = 20).

Tables 1-4 summarize the results for selected pointwise features of the incubation density for all scenarios (S1-S16). In general, the bias is relatively small for all features but is more pronounced for the 95*th* percentile as less information is available in that region in the sense that less data points are collected in such a remote location of the domain of the incubation density. In addition, we observe that an increase in the sample size leads to a decrease in the root mean square error (RMSE). This decrease comes sometimes at the cost of an increase in bias, reflecting the well-known bias-variance trade-off.

**Table 1:** Perfomance measures for selected features of the incubation density for two levels of data coarseness with sample size *n* = 100 and *n* = 200. Results are for *S* = 1000 simulated datasets with the lognormal incubation density of Ferretti et al. (2020).

| Average coarseness: 1 day |  |  |  |  |  |  |  |
| --- | --- | --- | --- | --- | --- | --- | --- |
| | True | $n = 100$ (S1) | | | $n = 200$ (S2) | | |
|  |  | Average | Bias | RMSE | Average | Bias | RMSE |
| Mean | 5.528 | 5.477 | -0.052 | 0.208 | 5.472 | -0.056 | 0.158 |
| SD | 2.075 | 1.993 | -0.082 | 0.195 | 1.997 | -0.078 | 0.154 |
| $q_{0.05}$ | 2.849 | 2.828 | -0.021 | 0.174 | 2.837 | -0.012 | 0.131 |
| $q_{0.25}$ | 4.052 | 4.053 | 0.001 | 0.171 | 4.047 | -0.005 | 0.124 |
| $q_{0.50}$ | 5.176 | 5.169 | -0.007 | 0.197 | 5.156 | -0.020 | 0.147 |
| $q_{0.75}$ | 6.612 | 6.559 | -0.053 | 0.262 | 6.546 | -0.066 | 0.203 |
| $q_{0.95}$ | 9.403 | 9.170 | -0.234 | 0.542 | 9.181 | -0.222 | 0.426 |
| Average coarseness: 2 days |  |  |  |  |  |  |  |
| | True | $n = 100$ (S3) | | | $n = 200$ (S4) | | |
|  |  | Average | Bias | RMSE | Average | Bias | RMSE |
| Mean | 5.528 | 5.431 | -0.097 | 0.225 | 5.417 | -0.111 | 0.186 |
| SD | 2.075 | 1.942 | -0.133 | 0.222 | 1.934 | -0.141 | 0.191 |
| $q_{0.05}$ | 2.849 | 2.836 | -0.013 | 0.179 | 2.847 | 0.002 | 0.134 |
| $q_{0.25}$ | 4.052 | 4.044 | -0.008 | 0.173 | 4.036 | -0.016 | 0.127 |
| $q_{0.50}$ | 5.176 | 5.138 | -0.038 | 0.205 | 5.118 | -0.057 | 0.158 |
| $q_{0.75}$ | 6.612 | 6.492 | -0.120 | 0.287 | 6.467 | -0.145 | 0.241 |
| $q_{0.95}$ | 9.403 | 9.022 | -0.381 | 0.624 | 9.002 | -0.401 | 0.536 |

**Table 2:** Perfomance measures for selected features of the incubation density for two levels of data coarseness with *n* = 100 and *n* = 200. Results are for *S* = 1000 simulated datasets and the Weibull incubation density of Backer et al. (2020).

| Average coarseness: 1 day |  |  |  |  |  |  |  |
| --- | --- | --- | --- | --- | --- | --- | --- |
| | True | $n = 100$ (S5) | | | $n = 200$ (S6) | | |
|  |  | Average | Bias | RMSE | Average | Bias | RMSE |
| Mean | 6.403 | 6.393 | -0.010 | 0.229 | 6.392 | -0.011 | 0.169 |
| SD | 2.327 | 2.325 | -0.002 | 0.151 | 2.327 | 0.001 | 0.110 |
| $q_{0.05}$ | 2.665 | 2.690 | 0.026 | 0.312 | 2.666 | 0.002 | 0.208 |
| $q_{0.25}$ | 4.734 | 4.726 | -0.008 | 0.262 | 4.722 | -0.012 | 0.191 |
| $q_{0.50}$ | 6.346 | 6.317 | -0.029 | 0.257 | 6.327 | -0.019 | 0.188 |
| $q_{0.75}$ | 7.995 | 7.961 | -0.034 | 0.270 | 7.975 | -0.020 | 0.199 |
| $q_{0.95}$ | 10.336 | 10.342 | 0.006 | 0.390 | 10.334 | -0.002 | 0.285 |
| Average coarseness: 2 days |  |  |  |  |  |  |  |
| | True | $n = 100$ (S7) | | | $n = 200$ (S8) | | |
|  |  | Average | Bias | RMSE | Average | Bias | RMSE |
| Mean | 6.403 | 6.350 | -0.053 | 0.232 | 6.338 | -0.065 | 0.181 |
| SD | 2.327 | 2.275 | -0.051 | 0.162 | 2.282 | -0.045 | 0.116 |
| $q_{0.05}$ | 2.665 | 2.712 | 0.048 | 0.308 | 2.666 | 0.002 | 0.202 |
| $q_{0.25}$ | 4.734 | 4.722 | -0.011 | 0.254 | 4.705 | -0.029 | 0.190 |
| $q_{0.50}$ | 6.346 | 6.282 | -0.064 | 0.256 | 6.282 | -0.064 | 0.198 |
| $q_{0.75}$ | 7.995 | 7.886 | -0.109 | 0.288 | 7.893 | -0.102 | 0.225 |
| $q_{0.95}$ | 10.336 | 10.202 | -0.133 | 0.434 | 10.187 | -0.149 | 0.321 |

**Table 3:** Perfomance measures for selected features of the incubation density for two levels of data coarseness with *n* = 100 and *n* = 200. Results are for *S* = 1000 simulated datasets and an artificial incubation density constructed as a mixture of two Weibulls.

| Average coarseness: 1 day |  |  |  |  |  |  |  |
| --- | --- | --- | --- | --- | --- | --- | --- |
| | True | $n = 100$ (S9) | | | $n = 200$ (S10) | | |
|  |  | Average | Bias | RMSE | Average | Bias | RMSE |
| Mean | 7.538 | 7.533 | -0.005 | 0.468 | 7.532 | -0.006 | 0.327 |
| SD | 4.622 | 4.593 | -0.029 | 0.143 | 4.599 | -0.023 | 0.098 |
| $q_{0.05}$ | 1.371 | 1.229 | -0.142 | 0.293 | 1.280 | -0.091 | 0.197 |
| $q_{0.25}$ | 3.050 | 3.026 | -0.024 | 0.311 | 3.005 | -0.045 | 0.207 |
| $q_{0.50}$ | 7.191 | 7.333 | 0.142 | 1.822 | 7.238 | 0.092 | 1.561 |
| $q_{0.75}$ | 12.080 | 12.027 | -0.053 | 0.315 | 12.023 | -0.057 | 0.215 |
| $q_{0.95}$ | 13.734 | 13.584 | -0.150 | 0.291 | 13.594 | -0.140 | 0.225 |
| Average coarseness: 2 days |  |  |  |  |  |  |  |
| | True | $n = 100$ (S11) | | | $n = 200$ (S12) | | |
|  |  | Average | Bias | RMSE | Average | Bias | RMSE |
| Mean | 7.538 | 7.493 | -0.045 | 0.486 | 7.506 | -0.032 | 0.328 |
| SD | 4.622 | 4.565 | -0.057 | 0.155 | 4.578 | -0.044 | 0.110 |
| $q_{0.05}$ | 1.371 | 1.228 | -0.143 | 0.297 | 1.282 | -0.089 | 0.196 |
| $q_{0.25}$ | 3.050 | 3.017 | -0.033 | 0.310 | 2.996 | -0.054 | 0.210 |
| $q_{0.50}$ | 7.191 | 7.275 | 0.084 | 1.940 | 7.248 | 0.057 | 1.627 |
| $q_{0.75}$ | 12.080 | 11.999 | -0.081 | 0.315 | 12.005 | -0.075 | 0.219 |
| $q_{0.95}$ | 13.734 | 13.376 | -0.358 | 0.445 | 13.407 | -0.327 | 0.377 |

From Figures 2-5, we see that in general the estimates provided by our method are able to nicely capture the target incubation densities. Thanks to the flexibility of our approach, even bimodal densities (Figure 4) are well reconstructed which would not be feasible with parametric approaches relying on classic families. Moroever, the dashed curves (representing the pointwise median of the estimates across the *S* = 1000 simulated datasets) are in most cases not distinguishable from the target incubation density. Also, the fitted densities appear closer to the target with *n* = 200 as compared to *n* = 100 as more information is available. This is corroborated by the squared Hellinger distance (see the histograms provided in the GitHub repository associated to this working paper) between the target incubation density *φ*(*·*) and our estimate *φ*(*·*) computed with the formula:

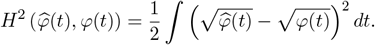

**Figure 2.**
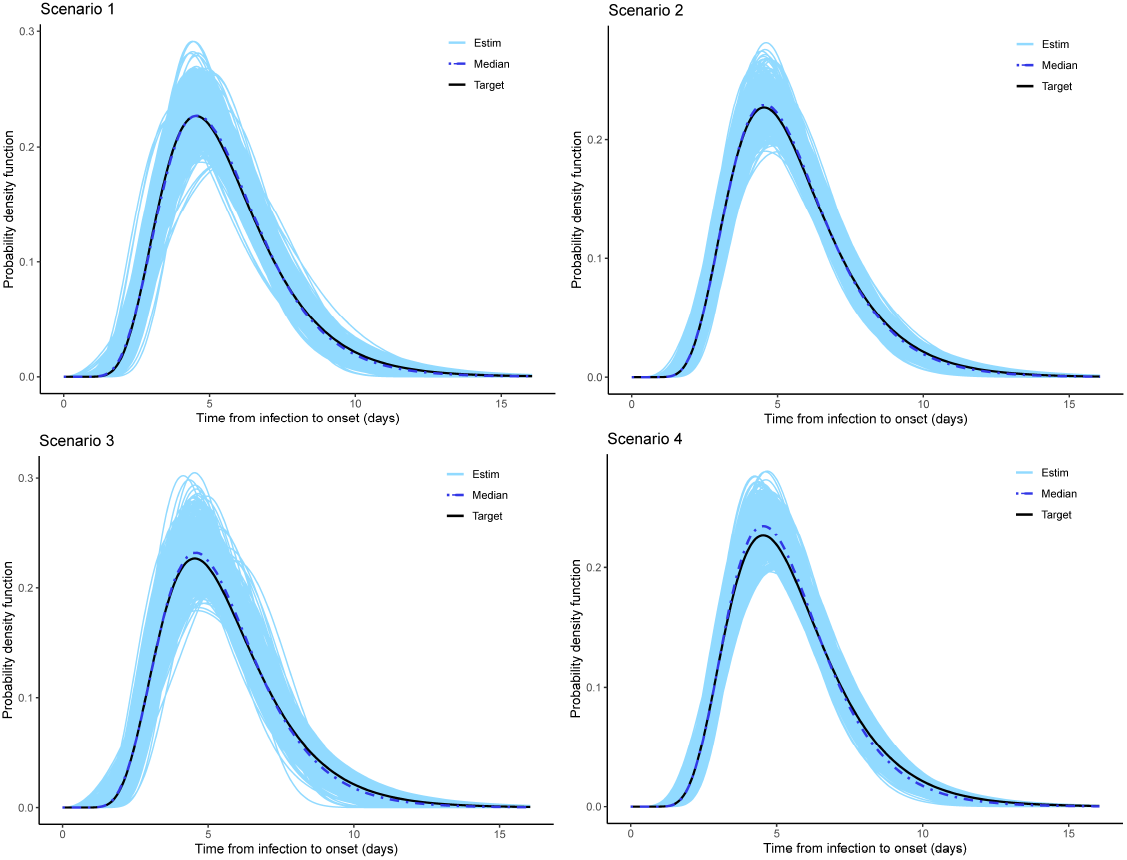
Estimated incubation densities for Scenarios 1-4. The dash-dotted line is the pointwise median across the S=1000 simulations and the solid black line is the lognormal incubation density of Ferretti et al. (2020).

**Figure 3.**
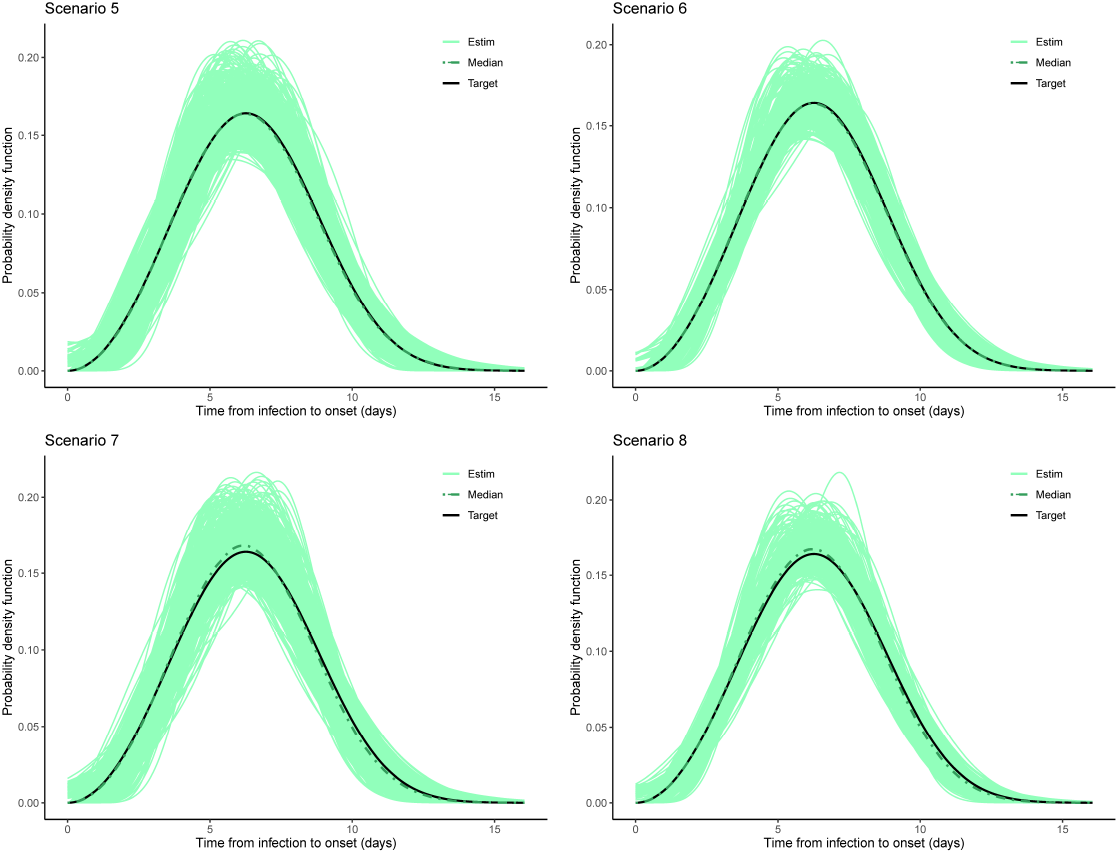
Estimated incubation densities for Scenarios 5-8. The dash-dotted line is the pointwise median across the S=1000 simulations and the solid black line is the Weibull incubation density of Backer et al. (2020).

**Figure 4.**
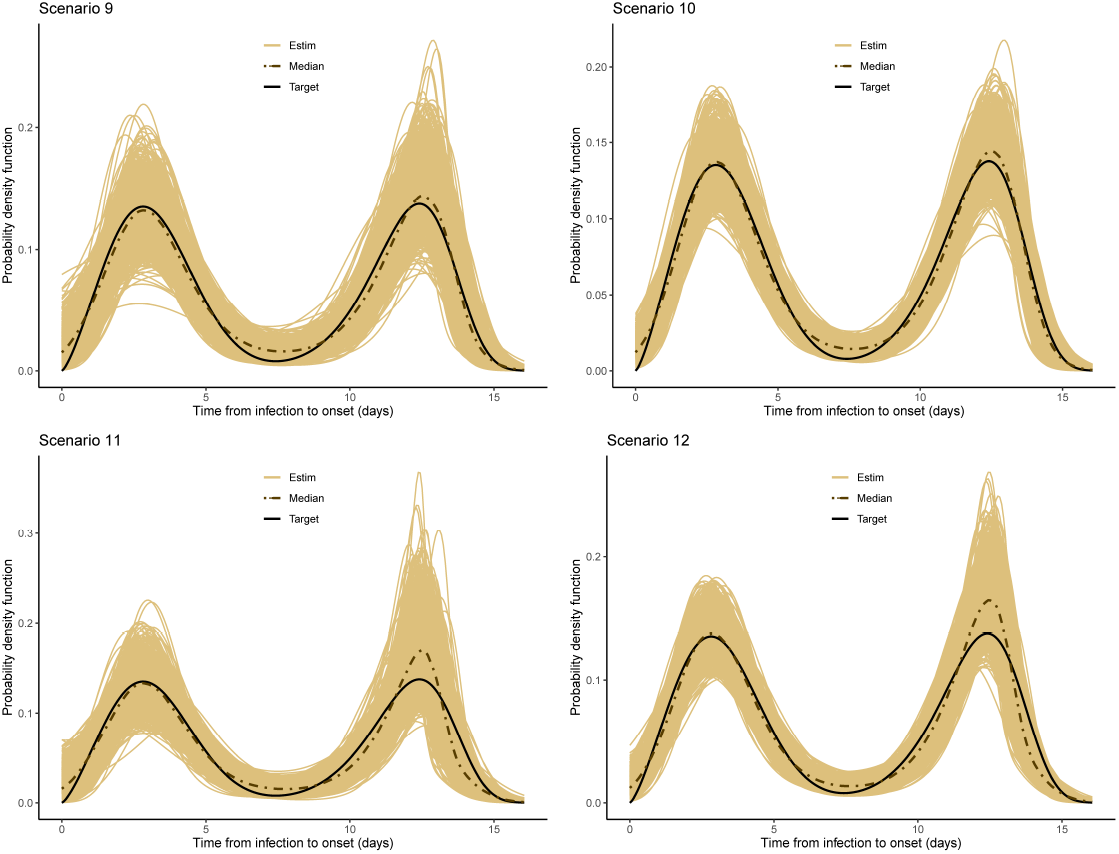
Estimated incubation densities for Scenarios 9-12. The dash-dotted line is the pointwise median across the S=1000 simulations and the solid black line is an artificial incubation density constructed as a mixture of two Weibulls.

**Figure 5.**
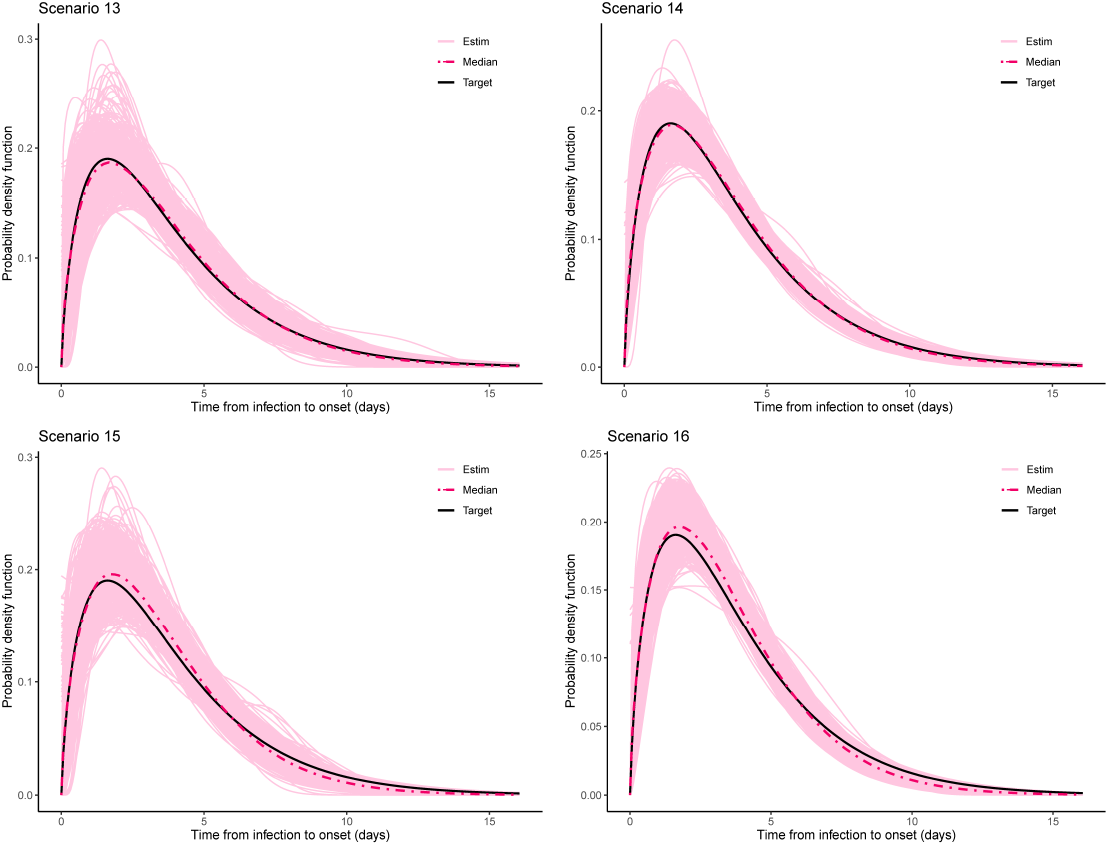
Estimated incubation densities for Scenarios 13-16. The dash-dotted line is the pointwise median across the S=1000 simulations and the solid black line is the Gamma incubation density of Donnelly et al. (2003).

**Figure 6.**
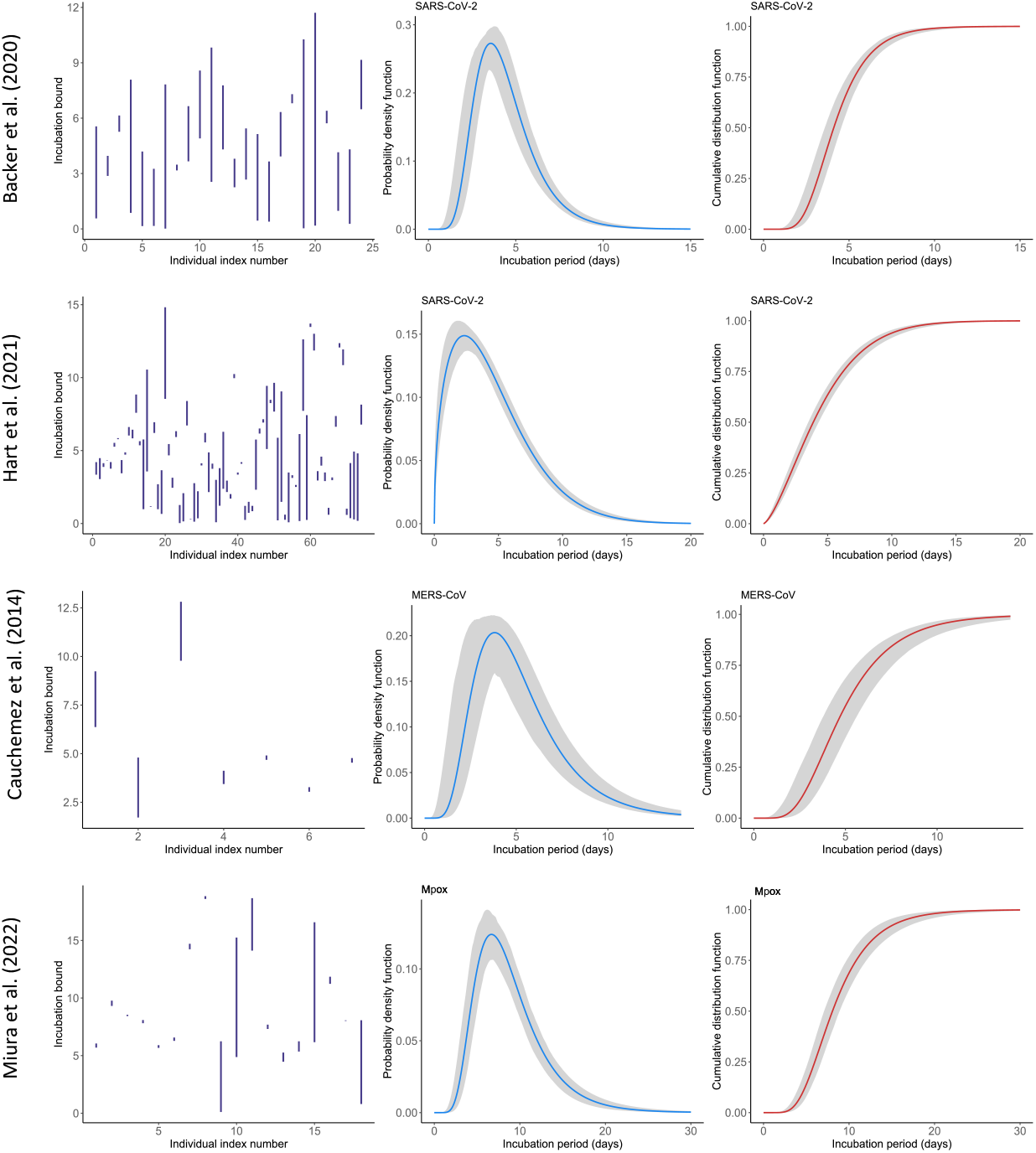
Incubation bounds and estimated probability density functions and cumulative distribution functions with the proposed flexible Bayesian approach for data on COVID-19, MERS-CoV and Mpox.

Finally, Table 5 shows that our method is quite efficient in detecting the true underlying distribution from which data is generated. For the lognormal incubation target, our LPS model selects the lognormal model in more than 73% of cases with *n* = 100 and it goes up to more than 80% of cases with *n* = 200. A correct selection is even made in 95% of cases in the Weibull setting. Interestingly, our methodology never selects any parametric candidate when the underlying truth is a bimodal density. Although this may not be the case for lower sample sizes, it is still an enouraging sign. Finally, for the Gamma case, our model hesitates between a Gamma and a Weibull but this is not really a problem as the main features of the true underling Gamma density are still relatively well captured (see Table 4).

**Table 4:** Perfomance measures for selected features of the incubation density for two levels of data coarseness with sample size *n* = 100 and *n* = 200. Results are for *S* = 1000 simulated datasets and the Gamma incubation density of Donnelly et al. (2003).

| Average coarseness: 1 day |  |  |  |  |  |  |  |
| --- | --- | --- | --- | --- | --- | --- | --- |
| | True | $n = 100$ (S13) | | | $n = 200$ (S14) | | |
|  |  | Average | Bias | RMSE | Average | Bias | RMSE |
| Mean | 3.810 | 3.756 | -0.054 | 0.298 | 3.738 | -0.072 | 0.205 |
| SD | 2.889 | 2.737 | -0.151 | 0.308 | 2.745 | -0.144 | 0.237 |
| $q_{0.05}$ | 0.561 | 0.564 | 0.003 | 0.134 | 0.554 | -0.008 | 0.091 |
| $q_{0.25}$ | 1.693 | 1.721 | 0.028 | 0.209 | 1.699 | 0.006 | 0.136 |
| $q_{0.50}$ | 3.110 | 3.135 | 0.025 | 0.288 | 3.109 | -0.001 | 0.189 |
| $q_{0.75}$ | 5.175 | 5.125 | -0.050 | 0.414 | 5.105 | -0.070 | 0.282 |
| $q_{0.95}$ | 9.451 | 9.063 | -0.388 | 0.867 | 9.073 | -0.377 | 0.649 |
| Average coarseness: 2 days |  |  |  |  |  |  |  |
| | True | $n = 100$ (S15) | | | $n = 200$ (S16) | | |
|  |  | Average | Bias | RMSE | Average | Bias | RMSE |
| Mean | 3.810 | 3.530 | -0.280 | 0.375 | 3.528 | -0.282 | 0.333 |
| SD | 2.889 | 2.462 | -0.426 | 0.485 | 2.472 | -0.417 | 0.449 |
| $q_{0.05}$ | 0.561 | 0.561 | 0.000 | 0.130 | 0.560 | -0.002 | 0.089 |
| $q_{0.25}$ | 1.693 | 1.681 | -0.012 | 0.186 | 1.675 | -0.018 | 0.131 |
| $q_{0.50}$ | 3.110 | 3.008 | -0.102 | 0.270 | 3.001 | -0.109 | 0.210 |
| $q_{0.75}$ | 5.175 | 4.820 | -0.355 | 0.503 | 4.818 | -0.357 | 0.438 |
| $q_{0.95}$ | 9.451 | 8.272 | -1.179 | 1.350 | 8.300 | -1.151 | 1.244 |

**Table 5:** Proportion of selected models across *S* = 1000 simulations under different scenarios.

| | $n = 100$ | | | | $n = 200$ | | | |
| --- | --- | --- | --- | --- | --- | --- | --- | --- |
| $\varphi_I \sim \mathbf{Lognormal}$ | SP | LN | G | W | SP | LN | G | W |
| (1 day coarseness) | 0% | 73% | 26.5% | 0.5% | 0% | 81.6% | 18.4% | 0% |
| (2 day coarseness) | 0% | 74.3% | 23.7% | 2% | 0% | 79.4% | 20.6% | 0% |
| $\varphi_I \sim \mathbf{Weibull}$ | $n = 100$ | | | | $n = 200$ | | | |
| (1 day coarseness) | 3.9% | 0.2% | 9.1% | 86.8% | 1.8% | 0% | 3.2% | 95% |
| (2 day coarseness) | 4.5% | 0.2% | 9.4% | 85.9% | 3.6% | 0% | 2.6% | 93.8% |
| $\varphi_I \sim \mathbf{Weibmix}$ | $n = 100$ | | | | $n = 200$ | | | |
| (1 day coarseness) | 100% | 0% | 0% | 0% | 100% | 0% | 0% | 0% |
| (2 day coarseness) | 100% | 0% | 0% | 0% | 100% | 0% | 0% | 0% |
| $\varphi_I \sim \mathbf{Gamma}$ | $n = 100$ | | | | $n = 200$ | | | |
| (1 day coarseness) | 2.7% | 1.2% | 58.1% | 38% | 0.5% | 0.1% | 62.8% | 36.6% |
| (2 day coarseness) | 3.9% | 0% | 42.6% | 52.9% | 0.5% | 0% | 42.8% | 56.7% |

**Table 6:** Description of the parametric distributions used in the moment matching approach.

| **Lognormal distribution** |  |
| --- | --- |
| Notation | $X \sim \text{LogNorm}(\alpha, \beta^2)$ |
| Parameters | $\alpha \in \mathbb{R}$ location; $\beta > 0$ scale |
| Density function | $p(x) = \frac{1}{x\sqrt{2\pi\beta^2}} \exp\left(-\frac{1}{2}\left(\frac{\log(x)-\alpha}{\beta}\right)^2\right)$ |
| Support | $x > 0$ |
| 1<sup>st</sup> moment (Mean) | $E(X) = \exp\left(\alpha + \frac{\beta^2}{2}\right)$ |
| 2<sup>nd</sup> central moment (Variance) | $V(X) = \exp(2\alpha + \beta^2) (\exp(\beta^2) - 1)$ |
| Moment matching | Root finding algorithm |
| **Gamma distribution** |  |
| Notation | $X \sim \mathcal{G}(\alpha, \beta)$ |
| Parameters | $\alpha > 0$ shape; $\beta > 0$ rate |
| Density function | $p(x) = \frac{\beta^\alpha}{\Gamma(\alpha)} x^{\alpha-1} \exp(-\beta x)$ |
| Support | $x > 0$ |
| 1<sup>st</sup> moment (Mean) | $E(X) = \frac{\alpha}{\beta}$ |
| 2<sup>nd</sup> central moment (Variance) | $V(X) = \frac{\alpha}{\beta^2}$ |
| Moment matching | Analytically available |
| **Weibull distribution** |  |
| Notation | $X \sim \text{Weibull}(\alpha, \beta)$ |
| Parameters | $\alpha > 0$ shape; $\beta > 0$ scale |
| Density function | $p(x) = \frac{\alpha}{\beta^\alpha} x^{\alpha-1} \exp\left(-\left(\frac{x}{\beta}\right)^\alpha\right)$ |
| Support | $x > 0$ |
| 1<sup>st</sup> moment (Mean) | $E(X) = \beta \Gamma\left(1 + \frac{1}{\alpha}\right)$ |
| 2<sup>nd</sup> central moment (Variance) | $V(X) = \beta^2 \left(\Gamma\left(1 + \frac{2}{\alpha}\right) - \left(\Gamma\left(1 + \frac{1}{\alpha}\right)\right)^2\right)$ |
| Moment matching | Root finding algorithm |

## 4 Applications to real data

This section applies the proposed flexible estimation methodology to publicly available datasets on reported exposures and symptom onset times. For the analyses, we use *K* = 20 B-splines and a MCMC chain of length *M* = 20, 000. A smart choice for *t*_*u*_ (and hence 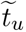), i.e. the upper bound on which to fix the B-spline basis can for instance be based on information from previous studies on the incubation period for a given pathogen. For instance, Virlogeux et al. (2015) reports the 99th percentile and range of the incubation period of human avian influenza A (H7N9) and the systematic review of Lessler et al. (2009) on incubation periods of acute respiratory viral infections gives an idea of the range of the incubation period for different diseases. Such empirical knowledge can help in finding a choice for *t*_*u*_ that supports with high confidence most of the probability mass of the incubation period distribution.

Another practical aspect worth mentioning is that exposure times and symptom onset times are in practice reported at a daily time resolution (calendar dates), while our model is in continuous time. A common strategy to transit from discrete to continuous observations is to assume that exact times are uniformly distributed throughout the day and hence to perturb symptom onset times and exposure window bounds by a uniform random variable between 0 and 1 (see e.g. Kreiss and Van Keilegom, 2022).

### 4.1 COVID-19 infections among travellers from Wuhan

First, we attempt to estimate the incubation density based on exposure times and symptom onset dates of confirmed COVID-19 cases with travel history to Wuhan (Backer et al., 2020). The analysis considers 25 visitors to Wuhan with a closed exposure window from which we removed an individual who had a quite large exposure period (20 days) as compared to the remaining observations. Backer et al. (2020) obtained a lognormal fit with a mean incubation period of 4.5 days (CI95%: 3.7-5.6) and a 95th percentile of 8.0 days (CI95%: 6.3-11.8). From a discussion with the first author of the latter reference, we were informed that a Gamma density with a mean of 4.6 days (CI95%: 3.8-5.4) and a 95th percentile of 7.4 days (CI95%: 6.2-9.7) fitted equally well. Our methodology provides a similar fit, namely a Gamma density with mean 4.4 days (CI95%: 4.0-4.8) and a 95th percentile of 7.7 days (CI95%: 7.2-8.5).

### 4.2 Transmission pair data on COVID-19

Next, we consider a dataset on transmission pairs for COVID-19 from Hart et al. (2021) that was analyzed (among others) in Xia et al. (2020). The latter reference obtained a Weibull fit for the incubation density with a mean of 4.9 days (CI95%: 4.4-5.4) and a 95th percentile of 9.9 days (CI95%: 8.9-11.2). Restricting our analysis to a subset of *n* = 74 individuals with closed exposure windows, we obtain a Weibull with a mean of 4.5 days (CI95%: 4.2-4.9) and a 95th percentile of 10.5 days (CI95%: 9.8-11.4).

### 4.3 Middle East Respiratory Syndrome (MERS)

In a third application, we consider a dataset given in Cauchemez et al. (2014) that reports lower and upper bounds of the incubation period for seven individual MERS-CoV cases in the United Kindom, France, Italy and Tunisia. Based on this data, the latter reference obtains a best fit to the incubation density that is lognormal with a mean of 5.5 days (CI95%: 3.6-10.2) and a 95th percentile of 10.2 days, extrapolated from the reported standard deviation in the reference (CI95%: NA). Our approach selects the semi-parametric fit with a mean of 5.3 days (CI95%: 4.5-6.2) and a 95th percentile of 10.1 days (CI95%: 9.2-12.1).

### 4.4 Mpox

The last application is on a dataset reporting *n* = 18 confirmed Mpox cases in the Netherlands (Miura et al., 2022). The latter analysis uses a parametric Bayesian approach similar to Backer et al. (2020) and the best fitting model is given by a lognormal distribution with a mean incubation period of 9.0 days (CI95%: 6.6-10.9) and a 95th percentile of 17.3 days (CI95%: 13.0-29.0). Analyzing the same dataset with our flexible Bayesian approach, we obtain a lognormal fit with mean incubation period of 8.9 days (CI95%: 7.9-9.9) and a 95th percentile of 16.6 days (CI95%: 14.7-19.1).

## 5 Discussion

This article proposes a flexible approach to tackle the challenging problem of estimating the incubation period distribution based on coarse data. This is done through Bayesian P-splines and Laplace approximations. The semi-parametric model approximates the incubation density via a finite mixture density smoother and the latter is used to fit three popular parametric distributions that are often considered in the esitmation of incubation times. The Bayesian infromation criterion is then able to arbitrate between the competing density estimators. Our methodology has a natural place in the EpiLPS ecosystem as density smoothing under imputation methods can be formulated as a histogram smoothing problem; a problem that has already been addressed by EpiLPS in the context of smoothing case incidence data to estimate the time-varying reproduction number. The current methodology also borrows from the exisiting Langevinized Gibbs sampler in EpiLPS and thus incubation estimation as proposed here has a natural nest in the EpiLPS package.

The main strength of our work is that it permits to go beyond classic parametric Bayesian approaches that are often considered in the literature. A clear disadvantage of such approaches is that they may miss important features of the incubation period distribution if the latter turns out to have a more flexible structure than what is proposed by parametric models. In addition, the methodology developed here does not close the door to classic parametric models as the latter can still be good candidates to get information on the incubation period. Another advantage is that the algorithms underlying our work are available in the EpiLPS package. As such, it can be of direct practical use for the scientific community and public health officers to analyze real datasets. Note also that the routines underlying EpiLPS are really efficient as computationally expensive parts are treated with C++, so that results are typically obtained in a few seconds.

The main limitation of this work is that in some rare cases, our approach may select a flexible fit to the incubation density while in reality it should have chosen a candidate among the parametric models. As suggested by the simulation study, this arises more frequently when the sample size is small (less than 5% of cases) rather than with larger sample sizes. For instance, with *n* = 200, this mismatch only happend in less than 3.6% of cases. Note however that even with the small sample sizes considered in the real data applications, our estimates for the mean and standard deviation of the incubation period seem to be well aligned with results from previous studies.

From here, there are several research paths to explore. We can for instance enrich the current model by not only considering a two-component mixture in the semi-parametric approach, but rather a multi-component mixture with a multiple imputation approach. Furthermore, if one has good ideas to believe that infection times are more likely to appear at another location than the midpoint of the exposure window, we can easily adjust this in our model. Note also that here, we gave equal weights *ω* = 0.5 to the single-interval censored data and midpoint imputed data methods. If prior knowledge is availabe on more specific locations of infection times, weights can be adjusted accordingly. Another interesting research perspective is to handle estimation of the generation interval (GI), i.e. the time difference between the infection event of a primary case (infector) and of a secondary case (infectee). As we now have a flexible method for estimating the incubation distribution, we could work under a convolution setting to propose an estimator for the GI.

## Acknowledgments

We thank Jantien Backer and Jacco Wallinga from the National Institute for Public Health and the Environment (RIVM) for discussing their results on the incubation period estimation for COVID-19 based on confirmed cases with Wuhan travel history.

## Data availability

Simulation results and real data applications in this paper can be fully reproduced with the code available on the GitHub repository (https://github.com/oswaldogressani/Incubation) based on the EpiLPS package version 1.2.0.

## Appendix S1

Let us define the following quantities related to the left bound of the incubation period:

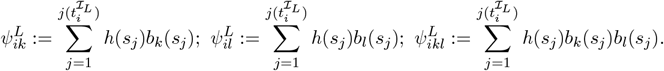

Analogously define the same quantities for the right bound 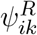, 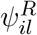 and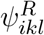.

### Gradient

Recall that the (approximated) log-likelihood is:

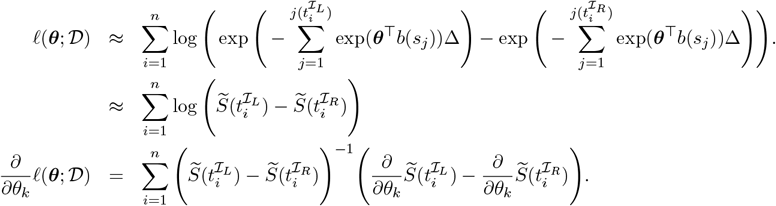

Note that:

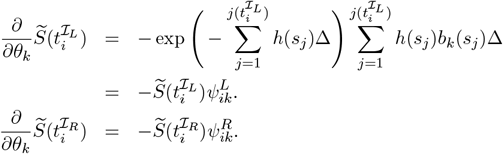

It follows that the *k*th entry to ∇_***θ***_*ℓ*(***θ***; *𝒟*) is:

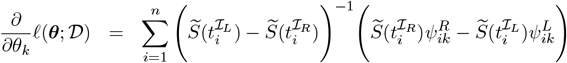

□

### Hessian

Let us define:

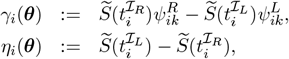

so that the *k*th entry to ∇_***θ***_*ℓ*(***θ***; *𝒟*) is rewritten compactly as:

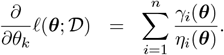

Deriving the above expression again with respect to the *l*th B-spline component gives:

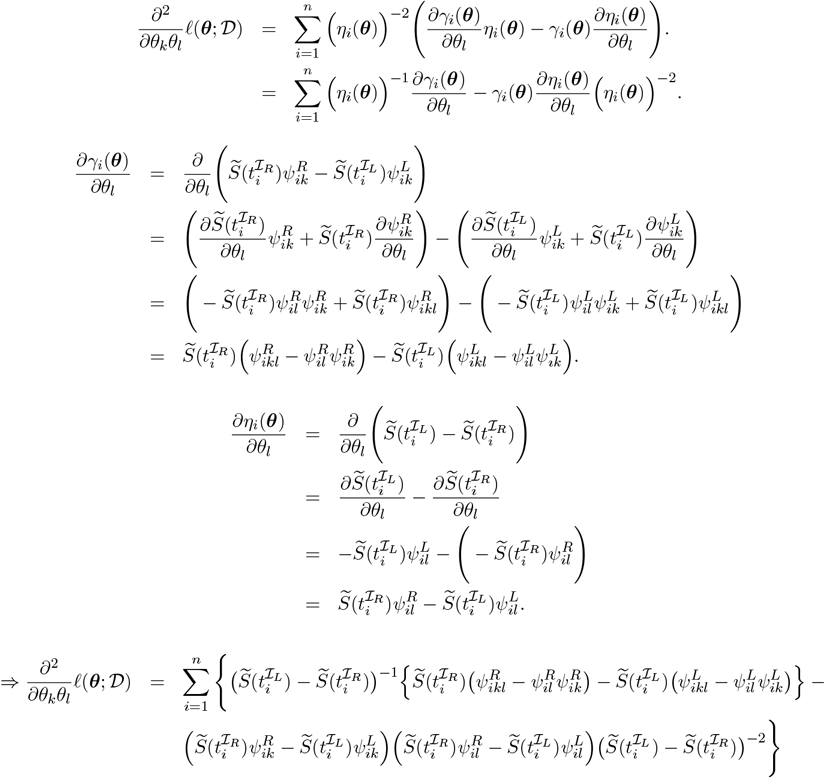

## Appendix S2

